# Detection of Hepatocellular Carcinoma from B-Mode and Contrast-Enhanced Ultrasound Using a Dual-Path Convolutional Network

**DOI:** 10.64898/2026.05.04.26352359

**Authors:** O. Francis, A. A. Roy

**Author notes:** Corresponding author: A. A. Roy.

## Abstract

**Background:** Hepatocellular carcinoma (HCC) is a leading cause of cancer-related mortality worldwide, with particularly severe consequences in sub-Saharan Africa where access to advanced diagnostic imaging remains limited. Ultrasound is the most widely available imaging modality in low-resource settings, yet its sensitivity for detecting early-stage HCC remains insufficient when used in conventional B-mode alone.

**Methods:** We present a dual-path convolutional neural network (CNN) that jointly analyzes B-mode and contrast-enhanced ultrasound (CEUS) images for automated HCC detection. The model processes 1,057 labeled liver ultrasound images from 85 patients sourced from The Cancer Imaging Archive, a publicly available single-center dataset. A preprocessing pipeline extracts liver-centered regions of interest from heterogeneous DICOM files, including automatic separation of dual-panel B-mode and CEUS frames. Each imaging modality is processed through a dedicated ResNet-34 backbone initialized with ImageNet weights, and the resulting feature embeddings are fused through a late-fusion classification head. The model is evaluated using patient-wise five-fold cross-validation and a held-out 20% patient-level test set.

**Results:** On the held-out test set, the model achieved 94.2% accuracy, 93.6% precision, 100% sensitivity, 83.3% specificity, and a 96.7% F1-score for binary HCC versus non-HCC classification. Cross-validation analysis showed consistently high discrimination across folds, with AUC values ranging from 0.93 to 0.98. Training dynamics indicated that early stopping typically activated between epochs seven and eleven, with validation loss closely tracking training loss and no evidence of severe overfitting under the chosen regularization scheme.

**Conclusions:** These findings demonstrate that a relatively lightweight multimodal CNN, trained on carefully preprocessed public data, can provide strong imaging-level discrimination between HCC and non-HCC findings within a single-center dataset. However, the small sample size, pronounced class imbalance, and single-center origin of the data preclude any claims of clinical utility at this stage. This work is a transparent, reproducible methodological baseline intended to support future multi-site validation, particularly in African and other low-resource clinical settings where ultrasound-based decision support could have the greatest impact.

## 1 Introduction

Liver cancer remains one of the most lethal malignancies worldwide, with hepatocellular carcinoma (HCC) accounting for more than 90% of primary liver tumors [Kwabena O and Hrishikesh, 2023, El-Kassas and Elbadry, 2022]. In 2022 alone, over 866,000 new cases and 758,000 deaths were recorded globally, and the highest age-standardized incidence and mortality rates were concentrated in East Asia and sub-Saharan Africa [Angucia et al., 2024]. Liver disease as a whole contributes more than 2 million deaths annually and represents approximately 4% of all global mortality [Devarbhavi et al., 2023]. In Africa, the rising burden is driven by an intersecting epidemic of chronic hepatitis B and C, alcohol misuse, aflatoxin B1 exposure, obesity, type-2 diabetes, and smoking [Tueller et al., 2023, El-Kassas and Elbadry, 2022]. Projections suggest the African liver cancer burden could double by 2030 because of population growth and persistent risk-factor exposure [Sung et al., 2021].

In developing countries, cancer incidence is rising rapidly. In Uganda, cancer incidence rose from 27,410 cases in 2012 to 35,968 in 2022, with cancer-related deaths increasing from 17,120 to 24,629 over the same period [Nakaganda et al., 2024]. The Uganda Cancer Institute receives 170 to 200 new HCC patients annually, the majority presenting at advanced stages ineligible for curative therapy [Nandudu, 2023]. This pattern of late diagnosis reflects both behavioral factors and severe structural constraints. Multiphasic computed tomography (CT) and magnetic resonance imaging (MRI) are available in only a handful of urban referral centers, and even when accessible, their cost, travel requirements, and long waiting times represent insurmountable barriers for rural patients.

Under these conditions, ultrasonography becomes the cornerstone of HCC surveillance and diagnosis in low- and middle-income countries (LMICs). B-mode ultrasound is portable, radiation-free, relatively inexpensive, and widely deployed even in district hospitals. However, its sensitivity for early-stage HCC in cirrhotic livers is modest, commonly reported below 60% and as low as 45% for lesions smaller than 2 cm, owing to coarse parenchymal echotexture, operator dependency, and patient-related factors such as obesity and hepatic steatosis [Frenette et al., 2019, Hanna et al., 2016]. Contrast-enhanced ultrasound (CEUS) dramatically improves lesion characterization by revealing arterial-phase hyperenhancement and late-phase washout typical of HCC, achieving sensitivity around 90% and specificity exceeding 97% in experienced hands [Tanaka, 2020]. Despite these advantages, CEUS uptake remains limited in sub-Saharan Africa by contrast-agent cost, supply-chain fragility, and the need for dedicated training.

Over the past decade, deep learning has transformed medical image analysis by learning hierarchical feature representations directly from raw data [Naji et al., 2019, Yin et al., 2025]. In liver oncology, convolutional neural networks (CNNs) applied to CT and MRI have reached or surpassed expert-level performance in lesion segmentation, malignancy classification, and treatment-response prediction [Guo et al., 2024, Zhang et al., 2023]. Ultrasound applications have been slower to mature because of greater image heterogeneity, speckle noise, and the historical scarcity of large annotated datasets. Nevertheless, recent studies using B-mode or CEUS alone have achieved AUC values consistently above 0.90 on curated cohorts [Zhang et al., 2022, Han et al., 2023, Guo et al., 2024, Tiyarattanachai et al., 2021], and several have outperformed non-specialist radiologists in focal liver lesion detection and characterization [Chaiteerakij et al., 2024].

Despite this progress, three critical gaps persist. First, most ultrasound-based models treat B-mode and CEUS as separate modalities, discarding the complementary information that clinicians routinely integrate when both modalities are acquired during the same examination. Second, nearly all published work relies on proprietary single-institution datasets, hampering reproducibility and external validation. Third, computational complexity and deployment feasibility in low-resource environments where GPU-enabled workstations are rare are seldom addressed explicitly.

This paper directly responds to these gaps by presenting a dual-path convolutional neural network that jointly processes B-mode and CEUS images, trained and rigorously evaluated on a heterogeneous public dataset from The Cancer Imaging Archive. The architecture is intentionally lightweight, uses transfer learning, and is designed for inference on commodity hardware. We emphasize patient-wise splitting, transparent preprocessing, and conservative interpretation of results given the single-center origin of the data and pronounced class imbalance. Rather than claiming immediate clinical readiness, we position this work as a reproducible methodological baseline to accelerate future multi-site validation in African and other low-resource contexts.

## 2 Related Work

### 2.1 Epidemiology and Clinical Imaging Pathways

HCC typically arises on a background of cirrhosis, which is present in up to 90% of cases [Llovet et al., 2021]. Risk factors vary geographically: hepatitis B predominates in sub-Saharan Africa and East Asia, whereas alcohol and metabolic dysfunction-associated steatotic liver disease are increasingly important in high-income settings [Cano et al., 2025]. The Barcelona Clinic Liver Cancer (BCLC) staging system integrates tumor burden, liver function, and performance status to guide therapy [Llovet et al., 2021]. Accurate imaging is essential because noninvasive diagnosis is now accepted for characteristic lesions in at-risk patients.

In resource-rich environments, surveillance relies on six-monthly ultrasound with alpha-fetoprotein, followed by multiphasic CT or MRI. In many African countries, however, CT and MRI capacity is extremely limited, and ultrasound becomes the de facto diagnostic modality despite its well-documented limitations [Frenette et al., 2019]. CEUS bridges part of this gap by providing real-time microvascular perfusion imaging without radiation or nephrotoxicity [Claudon et al., 2013, Tanaka, 2020], yet its adoption remains constrained by cost and training requirements.

### 2.2 Deep Learning in Liver Imaging

Early computational approaches extracted handcrafted radiomic features and applied classical classifiers such as support vector machines and random forests [Yasaka et al., 2018, Kather et al., 2019]. Deep learning has largely replaced these methods. In CT and MRI, CNNs and encoder-decoder networks now achieve expert-level segmentation and classification [Guo et al., 2024, Yin et al., 2025]. Ultrasound presents greater challenges because of speckle, shadowing, and operator variability, but progress has been rapid. B-mode CNNs have successfully discriminated HCC from hemangioma and focal nodular hyperplasia [Zhang et al., 2022, Yamakawa et al., 2019], while CEUS studies using 2D/3D CNNs or recurrent architectures have modeled enhancement dynamics with AUC values frequently exceeding 0.90 [Han et al., 2023, Guo et al., 2024].

Multimodal and dual-path strategies are gaining traction in the broader literature. Han et al. [Han et al., 2023] proposed a multi-view privileged-information framework combining B-mode and CEUS and achieved 88.91% accuracy in complex lesion classification. Li et al. [Li et al., 2024] fused clinical variables with ultrasound radiomics, reporting AUCs above 0.91.

Hybrid CNN-LSTM models that process entire CEUS cine loops without manual frame selection have reached AUC 0.91 [Guo et al., 2024]. Large-scale systems using YOLOv5 on more than 26,000 images have achieved overall focal-liver-lesion detection rates of 84.8% and malignancy sensitivity and specificity of 97% [Chaiteerakij et al., 2024].

Despite these advances, three persistent limitations motivate the present work. Most ultrasound models remain single-modality despite the routine clinical acquisition of both B-mode and CEUS. Datasets are almost universally proprietary and single-center, preventing reproducibility and external validation. Finally, few studies explicitly address computational footprint and deployment feasibility in low-resource clinical environments where GPU infrastructure is scarce.

### 2.3 Comparison with Existing Multimodal Fusion Approaches

Dual-path networks with late fusion have been applied across multiple imaging domains, including mammography [Brehar et al., 2020], retinal imaging [Yang et al., 2020], and chest radiography [Wang et al., 2021]. In these settings, late fusion typically concatenates or averages modality-specific feature vectors before passing them to a shared classifier. Alternative strategies include early fusion, in which raw inputs are stacked as additional channels prior to feature extraction, and intermediate fusion, in which feature maps are merged at one or more points within the network hierarchy.

In liver ultrasound specifically, Guo et al. [Guo et al., 2018] developed a two-stage multi-view learning framework that combined B-mode and CEUS features at multiple levels, and Tiyarattanachai et al. [Tiyarattanachai et al., 2021] applied single-modality deep learning to B-mode liver ultrasound in a large clinical cohort. The present work does not claim architectural novelty over these established paradigms. Instead, its contribution lies in the combination of a transparent, publicly reproducible pipeline with patient-wise evaluation on a heterogeneous public dataset, a configuration that has not been previously reported for joint B-mode and CEUS HCC classification. This distinction is methodological rather than algorithmic: the value lies in enabling independent replication and providing a benchmark against which more sophisticated architectures can be compared.

### 2.4 Public Datasets and Reproducibility

Public CT benchmarks such as LiTS [Bilic et al., 2022] and 3DIRCADb have catalyzed reproducible segmentation research, but ultrasound datasets remain scarce. The Cancer Imaging Archive recently released a B-mode and CEUS liver collection acquired during clinical trials of loco-regional therapy [tci, 2024]. Although single-center and heavily skewed toward confirmed HCC, its heterogeneity in scanner models, acquisition parameters, and image formats approximates real-world practice far better than tightly controlled research protocols. This public resource provides an ideal testbed for transparent algorithm development and benchmarking.

### 2.5 Gaps Motivating the Present Study

The literature reveals a clear trajectory from handcrafted features to end-to-end deep learning, from single-modality to multimodal fusion, and from proprietary to public data. Yet critical gaps remain for translation to sub-Saharan Africa: systematic joint modeling of B-mode and CEUS on publicly available data, rigorous patient-wise evaluation on heterogeneous imaging collections, and lightweight architectures compatible with modest hardware. The current work directly addresses these gaps by introducing a dual-path CNN designed for low-resource deployment, trained and evaluated on the TCIA ultrasound collection with full code and preprocessing pipeline released publicly.

## 3 Materials and Methods

### 3.1 Study Design and Endpoint

We adopted a design science methodology in which a technical artifact is designed, implemented, and empirically evaluated in response to a specific clinical challenge. In this case, the challenge is distinguishing HCC from non-HCC findings on liver ultrasound, and the artifact is the dual-path classification model. The primary endpoint is image-level binary classification of HCC versus non-HCC. Because multiple images can originate from the same patient, all data splits are performed at the patient level to avoid optimistic performance estimates arising from information leakage.

The overall pipeline consists of four stages: dataset curation and labeling, preprocessing and panel separation, model training with patient-wise cross-validation, and held-out test evaluation.

### 3.2 Dataset and Labeling

We used the liver ultrasound collection from The Cancer Imaging Archive, which includes B-mode and CEUS examinations acquired during routine clinical care. Figure 1 shows a sample image from the dataset. After excluding corrupted or unreadable files and non-diagnostic frames, we retained 1,057 images from 85 unique patients. The images include both single-panel examinations and dual-panel layouts in which B-mode and CEUS appear side by side.

**Figure 1:**
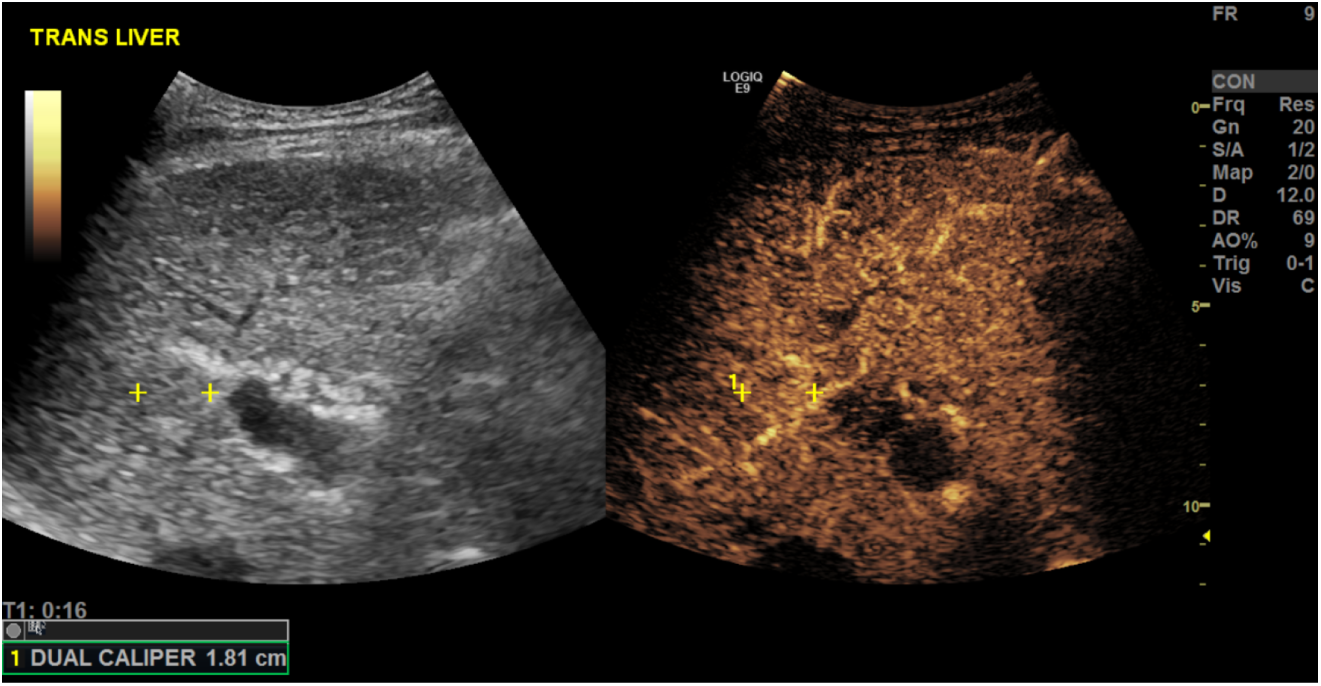
Example of a liver ultrasound image from the TCIA B-mode and CEUS Liver collection, showing a dual-panel layout with B-mode on the left and CEUS on the right.

Diagnostic labels were derived from radiology reports provided by the dataset publisher. Original labels comprised HCC, benign lesions, intrahepatic cholangiocarcinoma, indeterminate lesions, and composite categories. For this study we collapsed labels to a binary outcome: HCC versus non-HCC. Images from patients with histologically or radiologically confirmed HCC were assigned to the HCC class, and all remaining diagnoses, including cholangiocarcinoma and benign conditions, formed the non-HCC class. In the final dataset composition, HCC accounts for 935 images (88.5%), while non-HCC categories collectively contribute 122 images. This substantial class imbalance is an important consideration for both model training and the interpretation of evaluation metrics.

### 3.3 Preprocessing and Panel Separation

Raw DICOM files were downloaded using the NBIA Data Retriever and loaded in Python. The collection exhibited heterogeneity in spatial resolution, aspect ratio, and storage formats, including both single-frame images and cine loops. For multi-frame sequences, we selected a single representative frame based on metadata and frame indices, focusing on frames with clear depiction of the lesion and visible contrast phase when available.

Many examinations were stored as dual-panel images with B-mode and CEUS arranged horizontally. To separate these panels we computed the mean intensity along each image column and searched for a vertical band with low structural variation and characteristic separator appearance. The identified divider location was normalized by image width to maintain robustness across resolutions. Each frame was then split into left and right panels. A lightweight graphical review tool allowed manual correction of the divider for the minority of cases in which the automatic estimate was suboptimal.

Within each panel, a series of preprocessing steps were applied. Overlays such as text, calipers, and machine logos were detected using adaptive thresholding and morphological operations and removed by inpainting. The image was cropped to tight margins around the scan sector, and heuristic rules guided a coarse localization of the liver region. Morphological refinement produced a stable region of interest, which was then resized to a fixed input resolution with minimal distortion. Pixel intensities were standardized by subtracting the per-image mean and dividing by the per-image standard deviation. Figure 2 shows a sample B-mode image before and after preprocessing. Quality control rules excluded panels with near-uniform intensities or regions of interest below a minimum area threshold.

**Figure 2:**
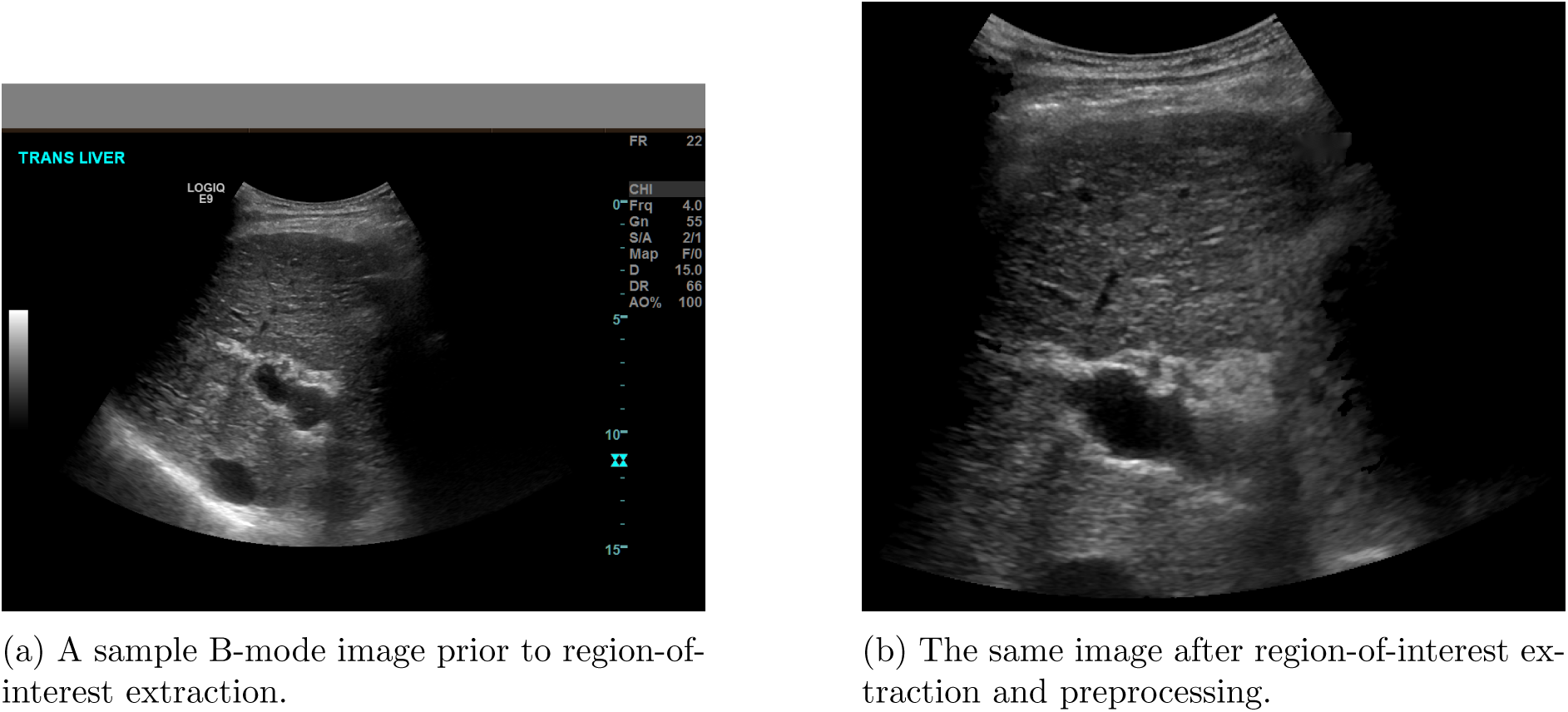
Illustration of the preprocessing pipeline showing a B-mode image before and after automated region-of-interest extraction. Overlays, calipers, and machine logos have been removed by adaptive thresholding and inpainting, and the image has been cropped to the liver parenchyma.

### 3.4 Train, Validation, and Test Splits

To prevent information leakage, data splits were defined at the patient level. All images from a given patient were allocated to the same subset. Patients were randomly partitioned into three disjoint groups: approximately 70% for training, 10% for validation via cross-validation, and 20% for a held-out test set. Splits were stratified to ensure that both HCC and non-HCC cases appeared in each subset.

Within the combined training and validation pool, we used five-fold cross-validation. In each fold, four-fifths of patients were used for training and one-fifth for validation. Hyperparameters and early stopping criteria were chosen based on validation performance aggregated across folds. After internal model selection, a final model was trained on the union of training and validation patients and evaluated once on the held-out test set.

### 3.5 Dual-Path Convolutional Neural Network

The classification model follows a dual-path design that mirrors clinical practice, in which B-mode and CEUS provide complementary perspectives on the same lesion. Each path uses a ResNet-34 backbone [He et al., 2016] truncated before the fully connected classifier and initialized with ImageNet [Deng et al., 2009] weights to leverage transfer learning. One backbone processes preprocessed B-mode regions of interest while the other processes CEUS regions.

Let *x*^(^*^B^*^)^ and *x*^(^*^C^*^)^ denote the B-mode and CEUS images, respectively. The respective back-bones compute feature vectors

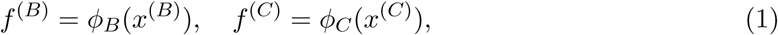

where *ϕ_B_* and *ϕ_C_* include convolutional layers and global average pooling. The resulting feature vectors are concatenated to form a joint embedding,

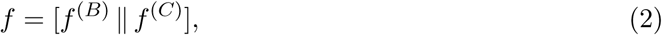

which is passed through a fusion head consisting of a fully connected layer, rectified linear unit activations, and dropout regularization. A final sigmoid neuron outputs the estimated probability that the image belongs to the HCC class.

A schematic of the architecture is shown in Figure 3.

**Figure 3:**
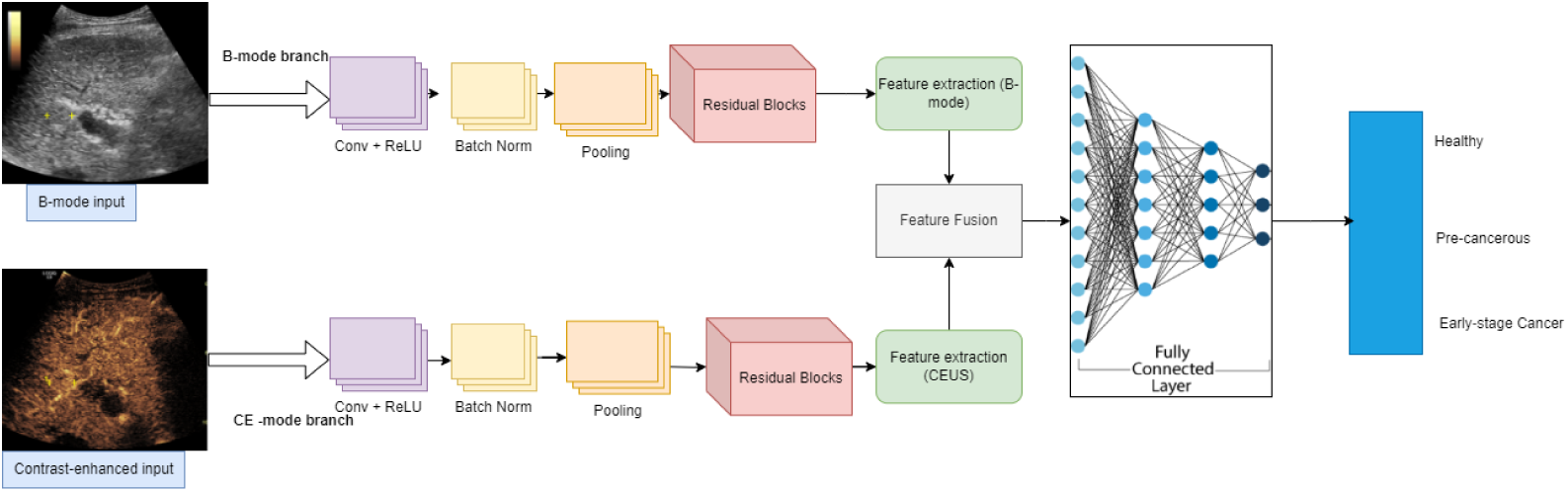
Dual-path convolutional neural network architecture. B-mode and CEUS images are processed by separate ResNet-34 backbones, and their feature embeddings are concatenated and passed through a fully connected fusion head to output the probability of HCC.

We emphasize that the dual-path late-fusion architecture is intentionally conventional. The goal of this study is not to propose a novel network topology but to establish a transparent, reproducible baseline using a well-understood design pattern on a public dataset. This deliberate choice allows future investigators to isolate the effects of more advanced fusion strategies, attention mechanisms, or transformer-based architectures by comparing directly against this baseline under identical data conditions.

### 3.6 Training Procedure and Data Augmentation

Training used the Adam optimizer with an initial learning rate of 10*^−^*^4^ and weight decay for regularization. Binary cross-entropy loss was minimized with class weights inversely proportional to class frequencies to mitigate the substantial imbalance between HCC and non-HCC categories. The learning rate was reduced on plateau according to validation loss, and early stopping with a patience of three epochs was used to prevent overfitting.

Online data augmentation was applied independently to B-mode and CEUS inputs. Augmentations included random horizontal flips, small rotations, scaling, mild elastic deformations to approximate probe pressure variation, and modest brightness and contrast adjustments. Augmentation ranges were chosen conservatively to reflect realistic image variability without generating implausible anatomies or enhancement patterns.

### 3.7 Evaluation Metrics

Model performance was quantified using accuracy, sensitivity, specificity, precision, F1-score, and area under the receiver operating characteristic curve (AUC-ROC). Confusion matrices were computed for each cross-validation fold and for the held-out test set. For the held-out test set we also report the Matthews correlation coefficient (MCC) as a summary of classification quality that accounts for all four entries of the confusion matrix and is therefore more informative than accuracy alone under class imbalance [Chicco and Jurman, 2020].

## 4 Results

### 4.1 Dataset Characterization

After preprocessing and quality control, the dataset comprised 1,057 images from 85 patients, with 935 images labeled as HCC and 122 as non-HCC. The number of images per patient varied, with a minority of patients contributing multiple frames in different contrast phases and the majority contributing a small number of key images. Both B-mode and CEUS panels were available for a large subset of examinations.

The strong dominance of HCC in both image and patient counts implies that naive classifiers could achieve high accuracy simply by predicting HCC for every input. Consequently, we emphasize sensitivity, specificity, F1-score, MCC, and AUC in both cross-validation and held-out evaluation rather than raw accuracy alone.

### 4.2 Training Dynamics

Figure 4 shows training and validation loss and accuracy for a representative cross-validation fold. Training and validation metrics improve rapidly over the first epochs and then plateau, with validation curves closely tracking training curves. Early stopping typically triggered between epochs seven and eleven across folds, suggesting that the learning rate and regularization were appropriately tuned. The close agreement between training and validation curves provides partial evidence against severe overfitting under the chosen configuration, although it does not rule out overfitting to the characteristics of this particular single-center dataset as a whole.

**Figure 4:**
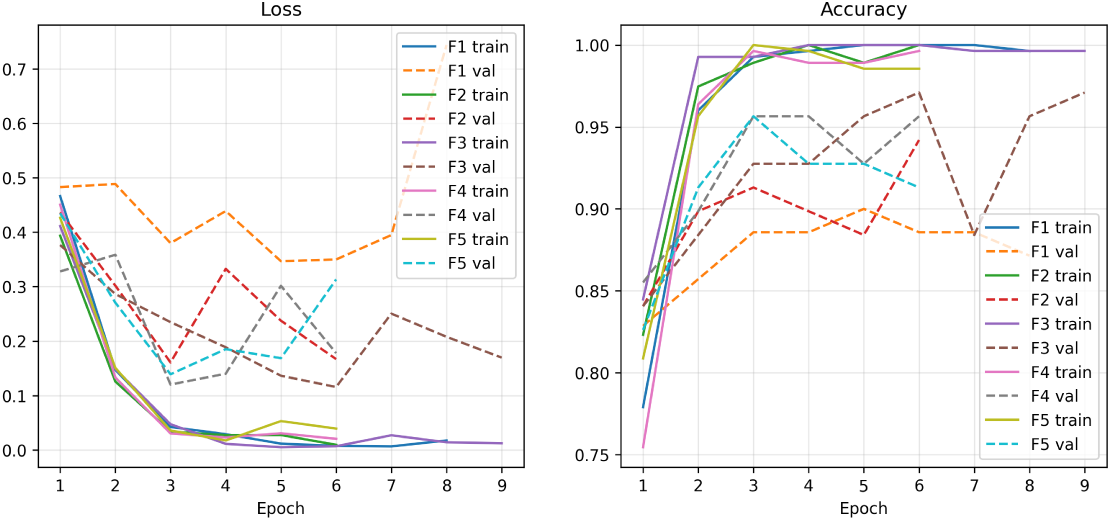
Training and validation loss curves and validation accuracy for a representative cross-validation fold, illustrating convergence behavior and the point at which early stopping activates.

### 4.3 Cross-Validation Performance

Table 1 summarizes patient-wise five-fold cross-validation performance. For each fold we report the total number of validation images, true positives, true negatives, false positives, false negatives, and AUC.

**Table 1:** Patient-wise five-fold cross-validation performance. TP: true positive (HCC correctly classified); TN: true negative (non-HCC correctly classified); FP: false positive; FN: false negative.

| Fold | Total | TP | TN | FP | FN | AUC |
| --- | --- | --- | --- | --- | --- | --- |
| F1 | 70 | 58 | 5 | 7 | 0 | 0.927 |
| F2 | 69 | 57 | 6 | 5 | 1 | 0.981 |
| F3 | 69 | 58 | 9 | 2 | 0 | 0.980 |
| F4 | 69 | 58 | 8 | 3 | 0 | 0.984 |
| F5 | 69 | 57 | 9 | 3 | 0 | 0.966 |

Sensitivity for HCC is high across folds, with only a single false negative observed in one fold out of five. Specificity is also strong, with non-HCC cases correctly recognized in most instances. AUC values range from 0.927 to 0.984, indicating excellent discrimination within this dataset. The limited variation in performance across folds suggests reasonable stability for different patient-wise partitions, although the small number of non-HCC patients per fold means that specificity estimates carry substantial sampling uncertainty.

Table 2 provides a summary of the aggregated cross-validation metrics alongside the held-out test set results, offering a compact comparison of internal and external evaluation.

**Table 2:** Aggregated model performance across cross-validation folds and on the held-out test set.

| Metric | CV Mean ± SD | Test |
| --- | --- | --- |
| Accuracy | $0.939 \pm 0.028$ | 0.943 |
| Precision | $0.936 \pm 0.027$ | 0.936 |
| Recall (Sensitivity) | $0.997 \pm 0.007$ | 1.000 |
| F1-Score | $0.965 \pm 0.016$ | 0.967 |
| AUC-ROC | $0.968 \pm 0.021$ | 1.000 |
| MCC | $0.763 \pm 0.115$ | 0.776 |

### 4.4 Held-Out Test Set Evaluation

After selecting the model configuration based on cross-validation, we retrained the model on the combined training and validation sets and evaluated it once on the held-out patient set. Table 3 presents the image-level performance metrics.

**Table 3:** Held-out test set performance for HCC versus non-HCC classification.

| Metric | Value |
| --- | --- |
| Accuracy | 94.2% |
| Precision | 93.6% |
| Recall (Sensitivity) | 100% |
| Specificity | 83.3% |
| F1-score | 96.7% |
| AUC-ROC | 1.00 |
| Matthews correlation coefficient | 0.775 |

The model attains 94.2% accuracy with perfect sensitivity for HCC and high precision. Specificity remains above 80%, indicating that most non-HCC images are correctly identified. The AUC of 1.00 on the held-out test set and the perfect sensitivity warrant careful interpretation. With only a small number of non-HCC patients in the test partition, these metrics are heavily influenced by individual cases and may not generalize. The MCC of 0.775 offers a more balanced assessment and confirms that the model performs substantially better than chance, but it also highlights that discrimination is not as strong as the headline AUC might suggest.

False positives mainly correspond to non-HCC lesions with imaging characteristics overlapping those of HCC, including hypervascular lesions in cirrhotic livers and cases with imaging artifacts. From a screening perspective, such false positives would be expected to undergo further diagnostic work-up, and their number is modest relative to the number of correctly identified HCC images.

### 4.5 Critical Assessment of Metrics

The near-perfect sensitivity and AUC on the held-out test set require explicit scrutiny, as such results are uncommon in clinical imaging studies and may signal favorable evaluation conditions rather than exceptional model performance. Several factors likely contribute to these optimistic estimates. The class imbalance is severe, with HCC comprising 88.5% of images, meaning that very few non-HCC samples are available to challenge the model. The test set, while patient-disjoint from training, is drawn from the same institution and acquisition protocols, eliminating the domain shift that typically degrades performance in external validation. Furthermore, the binary classification task collapses heterogeneous non-HCC diagnoses into a single category, which may be easier to discriminate from HCC than a more clinically realistic multi-class formulation would be.

We therefore caution readers against interpreting these results as indicative of deployment-ready performance. The metrics establish that the proposed pipeline is technically functional and capable of learning discriminative features from this specific dataset, but they do not constitute evidence of clinical validity. External validation on independent cohorts from different institutions, scanner vendors, and patient populations is essential before any stronger conclusions can be drawn.

## 5 Discussion

### 5.1 Principal Findings

We developed a dual-path CNN that processes B-mode and CEUS liver ultrasound images and evaluated it on a heterogeneous public dataset using patient-wise splits. The model achieved high sensitivity, specificity, and AUC in both cross-validation and held-out testing, with no missed HCC images in the held-out set. These results demonstrate that a relatively compact multimodal CNN, trained on carefully preprocessed data, can provide strong imaging-only discrimination between HCC and non-HCC within this single-center dataset.

### 5.2 Clinical Interpretation

From a clinical standpoint, the most important finding is the combination of high sensitivity and high F1-score. In surveillance or triage settings, missing a treatable HCC lesion carries substantial risk. The absence of false negatives on the held-out test set, together with consistently high sensitivity across cross-validation folds, suggests that the model tends to err on the side of flagging suspicious images rather than dismissing them. This behavior is desirable for a screening tool, where the cost of a missed malignancy far exceeds the cost of an unnecessary follow-up examination.

False positives, while undesirable, are generally less clinically harmful than false negatives in this context, since they can be resolved by human review and additional imaging where available. Examination of misclassified non-HCC images indicates that many represent challenging lesions or suboptimal image quality, which would likely also prompt careful review by human readers in routine clinical practice.

### 5.3 Value of Multimodal Fusion

The dual-path design directly encodes the clinical intuition that B-mode and CEUS are complementary modalities. B-mode images convey structural and textural information about the lesion and surrounding liver parenchyma, while CEUS highlights perfusion patterns including the arterial hyperenhancement and washout dynamics characteristic of HCC. By allowing each path to learn modality-specific features and then fusing their embeddings, the model can capture interactions between structure and enhancement without forcing a single modality to carry all diagnostically relevant information.

We did not perform exhaustive ablation experiments in this study, which represents a limitation. Formal comparison of the dual-path architecture against B-mode-only and CEUS-only baselines under identical training conditions is necessary to quantify the specific contribution of multimodal fusion. We plan to conduct these ablations in future work and recognize that without them, the added value of the dual-path design over single-modality alternatives remains an empirical hypothesis rather than a demonstrated finding.

### 5.4 Relation to Prior Work

The results align with and extend previous work applying deep learning to liver imaging [Guo et al., 2024, Zhang et al., 2023, Fang et al., 2021, Han et al., 2023, Yamakawa et al., 2019, Zhang et al., 2022]. Like earlier CT- and MRI-based models, our approach uses a CNN backbone with transfer learning to achieve high discrimination. Unlike many prior ultrasound studies, we explicitly integrate B-mode and CEUS in a single architecture and rely on a publicly accessible dataset, which facilitates reproducibility and benchmark comparisons.

It is important to acknowledge that the dual-path late-fusion architecture does not represent a methodological advance over existing multimodal fusion strategies in the literature. Late fusion of modality-specific feature vectors is a well-established paradigm, and more sophisticated alternatives including cross-modal attention, feature pyramid fusion, and transformer-based architectures have been proposed for related tasks [Han et al., 2023, Wang et al., 2024]. The contribution of this work is not architectural novelty but rather the transparent application of a known design pattern to a public dataset under rigorous patient-wise evaluation, providing a reproducible baseline that the community can build upon.

The architecture is intentionally modest in depth and parameter count compared with recent attention-based or transformer-inspired models. Achieving strong performance with such a compact network on this dataset suggests that careful preprocessing and sensible multimodal fusion can compensate, in part, for architectural complexity, and that such compact models may be more compatible with deployment on standard hospital hardware in resource-constrained settings.

### 5.5 Comparison with State-of-the-Art Methods

Direct quantitative comparison with existing methods is complicated by differences in datasets, label definitions, evaluation protocols, and the proprietary nature of most published cohorts. Nevertheless, contextualizing our results within the broader literature is essential for assessing relative performance.

Tiyarattanachai et al. [Tiyarattanachai et al., 2021] applied a CNN to B-mode liver ultrasound across a large multi-center cohort of over 44,000 images and reported an AUC of 0.92 for focal liver lesion detection. Their study benefits from far larger sample size and multi-center diversity but uses B-mode alone without CEUS integration. Han et al. [Han et al., 2023] achieved 88.91% accuracy using a multi-view framework combining B-mode and CEUS, a result that is lower in absolute terms than our 94.2% but was obtained on a more challenging multi-class classification task with a larger and more diverse patient cohort. Guo et al. [Guo et al., 2024] reported an AUC of 0.91 using a hybrid CNN-LSTM model on CEUS cine loops, leveraging temporal enhancement dynamics that our static-frame approach does not capture.

These comparisons illustrate that our numerical results fall within the range reported in the literature, but they do not permit claims of superiority. The differences in dataset composition, task formulation, and evaluation methodology are too substantial for fair head-to-head comparison. The primary advantage of our work relative to these studies is reproducibility: because both the data and the preprocessing pipeline are publicly available, independent investigators can replicate and extend the experiments, which is not possible for most published approaches.

### 5.6 Implications for Resource-Constrained Settings

Although the training data originate from a single institution that is not in sub-Saharan Africa, several aspects of the design have relevance for low-resource settings. The approach uses ultrasound, which is widely available, and relies only on post-acquisition DICOM exports, which can be obtained from many existing scanners without hardware modification. The model can run inference within seconds on a mid-range GPU or a modern CPU with optimized inference libraries, making real-time or near-real-time decision support plausible even without dedicated GPU infrastructure.

In a hypothetical deployment at an African regional referral hospital, the pipeline could operate as follows. Ultrasound examinations performed for chronic liver disease surveillance would be exported to a workstation running the preprocessing and inference software. The model would assign an HCC probability to each examination and highlight high-risk cases for review by a radiologist or hepatologist. This workflow does not remove the need for cross-sectional imaging or biopsy where indicated, but could help prioritize limited specialist time and advanced imaging resources.

However, given the single-center origin of the training data, direct transfer to LMIC settings is not guaranteed. Differences in ultrasound scanners, patient populations, disease prevalence, and acquisition protocols may substantially degrade performance. Domain adaptation strategies, local fine-tuning on African datasets, and prospective clinical validation studies would all be necessary before any deployment could be considered. This study should therefore be viewed as establishing a transparent baseline architecture and pipeline that can be retrained and recalibrated on local data, rather than as an off-the-shelf clinical solution.

### 5.7 Limitations

Several limitations merit explicit discussion and directly inform the scope of conclusions that can be drawn from this work.

First, the dataset is single-center. While heterogeneous in image format and quality, it does not capture the full variability across institutions, scanner vendors, and patient populations. External validation on independent cohorts from different regions and care levels is essential before considering clinical deployment. The absence of external validation is perhaps the most significant limitation of the current study.

Second, we consider a binary outcome of HCC versus non-HCC. For clinical decision-making, it may be important to distinguish specific non-HCC diagnoses, such as cholangiocarcinoma versus benign lesions, and to incorporate lesion size and stage. Extending the model to multi-class classification or to regression on clinically relevant risk scores is a logical next step.

Third, the model operates on static images or selected frames from cine loops. Full exploitation of CEUS temporal dynamics would require sequence-based architectures, such as 3D CNNs or hybrids with recurrent units. Temporal modeling may further improve performance, particularly for lesions with subtle enhancement patterns.

Fourth, our approach uses only imaging information. In practice, integrating laboratory data such as liver function tests and alpha-fetoprotein levels alongside clinical risk factors would likely yield better-calibrated risk estimates and align more closely with established HCC surveillance guidelines.

Fifth, although we used weighted loss functions and patient-wise splits, the class imbalance remains substantial. The held-out test set is relatively small, and performance estimates may be optimistic. Confidence intervals, bootstrapped performance estimates, and calibration analyses were not computed in this initial study, and their absence limits the statistical rigor of the reported metrics.

Sixth, no formal ablation study was conducted to isolate the contribution of the dual-path architecture relative to single-modality baselines. Without B-mode-only and CEUS-only comparisons under identical conditions, the specific value added by multimodal fusion cannot be conclusively established.

Finally, the near-perfect AUC and sensitivity on the held-out set, while encouraging, should be interpreted in light of the small test sample and single-center data provenance. These results establish technical feasibility but do not constitute evidence of clinical effectiveness.

### 5.8 Future Work

Future work should prioritize multi-site validation and local retraining on datasets from African and other LMIC institutions, ideally including both tertiary centers and regional hospitals. This would allow assessment of generalizability and identification of scanner-specific or population-specific failure modes. Prospective studies could evaluate the model in real clinical workflows, quantifying its impact on time to diagnosis, referral patterns, and downstream imaging utilization.

Methodologically, we plan to perform explicit ablation studies comparing single-modality and dual-path variants under identical training conditions. We also intend to explore temporal modeling of CEUS sequences using recurrent or 3D convolutional architectures, to investigate self-supervised or semi-supervised pretraining on large unlabeled ultrasound archives, and to evaluate more advanced fusion strategies including cross-modal attention mechanisms and feature pyramid networks. Comparison against recent transformer-based architectures for medical image analysis would further clarify whether the performance ceiling of the current approach reflects architectural limitations or data constraints.

From a deployment perspective, user interface design, visualization of saliency maps or attention regions to support clinical interpretability, and structured training programs for clinicians to interpret model outputs will be essential. Regulatory pathway considerations and ethical frameworks for AI-assisted diagnosis in LMIC settings also require careful attention. Ultimately, the most promising role for this type of model is as a human-in-the-loop decision support tool that augments rather than replaces clinical judgment.

## 6 Conclusion

We have presented a dual-path convolutional neural network that jointly processes B-mode and CEUS liver ultrasound images to distinguish HCC from non-HCC findings. Trained and evaluated on a heterogeneous single-center public dataset with patient-wise splits, the model achieves high sensitivity, specificity, and AUC, with no missed HCC images in the held-out test set. The architecture is compact and compatible with deployment on standard hospital hardware.

These results establish a reproducible methodological baseline for multimodal ultrasound-based HCC classification on publicly available data. The study does not claim clinical readiness, and the high performance metrics should be interpreted with caution given the small sample size, pronounced class imbalance, and single-center data origin. External validation on multi-site cohorts, formal ablation studies comparing fusion strategies, and prospective evaluation in clinical settings are necessary next steps. By releasing both the preprocessing pipeline and the model code alongside a public dataset, we aim to lower the barrier for future investigators to build upon this work, with the ultimate goal of developing validated ultrasound-based decision support tools for low-resource settings where they are needed most.

## Data Availability

The ultrasound images used in this study are publicly available through The Cancer Imaging Archive. The preprocessing pipeline and model training code will be made available upon request.

https://www.cancerimagingarchive.net/collection/b-mode-and-ceus-liver/

## Data and Code Availability

The ultrasound images used in this study are publicly available through The Cancer Imaging Archive. The preprocessing pipeline and model training code will be made available upon publication.

## Ethics Statement

This study used de-identified, publicly available data from The Cancer Imaging Archive. No patient contact or additional data collection was performed. Institutional review board approval was not required for analysis of this publicly released, de-identified dataset.

## Conflict of Interest

The authors declare no conflicts of interest.

## Acknowledgments

The authors acknowledge The Cancer Imaging Archive for making the liver ultrasound dataset publicly available.

## Notes

### Competing Interest Statement

The authors have declared no competing interest.

### Clinical Trial

IND126768

### Funding Statement

This study did not receive any funding

### Author Declarations

This study used de-identified, publicly available data from The Cancer Imaging Archive (https://www.cancerimagingarchive.net/collection/b-mode-and-ceus-liver/). No patient contact or additional data collection was performed. Institutional review board approval was not required for analysis of this publicly released, de-identified dataset.

